# Knowledge, attitude and utilization of nursing theories among nursing personnel: A descriptive cross-sectional study

**DOI:** 10.1101/2024.05.21.24307677

**Authors:** Joyce Joseph, Linnet Maria Tom, Krishna Maji, Kusumlata Sahu, Lakshmi Gupta, KM Akriti Bajpai, Komalpreet Kaur

**Affiliations:** All India Institute of Medical Sciences, Raipur; AIIMS, Raipur

**Keywords:** Nursing personnel, nursing theories, knowledge, attitude, utilization

## Abstract

**Aim:** To assess knowledge, attitude and utilization of Nursing Theories among Nursing Personnel working in a tertiary care centre in India.

**Methods:** A descriptive cross-sectional study was conducted using a proportionate stratified random sample of 350 nursing personnel in a tertiary care centre in east-central India. A self-structured tool to assess knowledge, practice and utilization was distributed to nursing personnel via Google forms between, March 2024-April 2024.

**Results:** The results revealed that the mean scores for the knowledge, attitude, and utilization component being 2.58 ± 0.570, 53.64 ± 6.433, and 1.87 ± 0.740, respectively. Among the nursing personnel, the analysis does not show any significant (p-value > 0.05) difference in the mean scores of knowledge, attitude and utilization when comparing different designations, age groups, genders, levels of work experience, professional qualifications, and types of institutes, indicating that the observed differences could be due to chance. However, in utilization, when examining the nature of the job, a significant difference is observed.

**Conclusions:** It is important to practice nursing theories in clinical setting for better patient care. Theories of nursing contribute to richness of the profession and provide an identity to the profession. Barriers in implementing nursing theories in practice must be examined and a particular focus to be given.

## Introduction

Nursing profession is not merely about administering injections or giving medications, but it’s all about holistically providing care. The prime reason for including nursing theories in nursing curriculum is to advance the discipline and professional practice of nursing. Even though different theories have different perspectives and meanings; they play an inevitable role in explaining the key notions and philosophy of nursing practice in an easy manner.^1^ When nursing theories are not put in practice, the contributions of nursing will become unclear, which makes the nurses just do the assigned tasks, that fall outside the scope of nursing practice.^2^

The present era emphasizes more on utilization of theoretical works to guide practice. Discipline specific knowledge; contribute to the safety and quality of care.^3^ Despite this importance, little attention is given to the practice of nursing theories among nurses in India as compared to other developed countries. There is a scarcity of literature available regarding the application of theory into practice in our Indian clinical setting. The primary responsibility for effectively implementing nursing theories in clinical settings lies with nursing institutions and researchers, aiming to enhance nursing practices and advance the profession.^4^

This study is the need of the hour as baseline knowledge, attitude, and practice of nursing theories of the target population should be obtained before planning any training programs or educational activities or policy changes. Very little data is available in the literature highlighting the knowledge, attitude, and utilization regarding nursing theories.

### Objectives of the study

- To assess the knowledge of nursing theories among nursing personnel in AIIMS Raipur.
- To determine the attitude of nursing theories among nursing personnel in AIIMS Raipur.
- To detect the utilization of nursing theories among nursing personnel in AIIMS Raipur.
- To find out the association between level of knowledge, attitude and utilization of nursing theories among nursing personnel and selected demographic variables.

## Methodology

The study was a descriptive cross-sectional study. The setting of the study was All India Institute of Medical Sciences, Raipur, Chhattisgarh, India. The data regarding the population was obtained from the administrative department.

### Study Sample

In this study target population consisted of Nursing Personnel working in All India Institute of Medical Sciences, Raipur, Chhattisgarh, which included Nursing Officers (NO), Senior Nursing Officers (SNO), Assistant Nursing Superintendent (ANS), Deputy Nursing Superintendent (DNS) and Nursing Tutors. As the population size was finite and known, Slovin’s formula was used to determine the sample size of 350 from a population size of 1580 and margin of error 0.05. Proportionate stratified random sampling done from the accessible population resulted in 266 Nursing Officers, 66, Senior Nursing Officers, 7 Assistant Nursing Superintendents, 4 Deputy Nursing Superintendents and 7 Nursing Tutors. The inclusion criterion was nursing personnel working on permanent basis in the institute. The exclusion criterion was those nursing personnel having done their diploma course in General Nursing and Midwifery. The participants were informed about the purpose and significance of the study, as well as the confidentiality and voluntary nature of their participation. The main study was conducted in the months of March and April using Google form for data collection. There was a little bit of reluctance to participate from DNS and Nursing Officers listening to the topic of the study.

### Ethical Consideration

- Prior permission was obtained from Research Review Committee (RRC). CON/AIIMSRPR/RRCN/2023/05
- Prior permission was obtained from the Institutional Ethical Committee (IEC). 4273/IEC-AIIMSRPR/2024
- Prior permission was obtained from AIIMS Authorities.
- Informed written consent was taken prior to the study from the study subjects.

### Study Tool and Data Collection

The study employed socio-demographic information form and a self-structured tool with four parts, Tool-1 was structured knowledge questionnaire with 10 questions in which correct answer was coded as 1 and wrong answer as zero (0). Tool-2 a five-point likert scale with 14 questions (Strongly Disagree, Disagree, Neutral, Agree, and Strongly Agree). Where strongly agree was coded as 5, agree as 4, neutral as 3, disagree as 2, and strongly disagree as 1 except for question11, which was reverse scored. Tool-3 was a structured practice checklist with 10 questions, each of which was scored on a two-level scale of Yes or No, in which Yes answer was coded as 1 and No answer as zero (0). The tools were developed by extensive review of literature, and item analysis was done. The content validity was done by 7 experts in the field, which obtained a content validity index (CVI) of 0.82. Test-retest reliability was done using Karl-Pearson Correlation Coefficient (0.78) for Tool-1. Internal consistency was established using Cronback’s alpha (0.82) for Tool-2 and Kuder Richardson-20 (0.90). The tools thus demonstrated satisfactory reliability and validity. Pilot study was done in February and the main study during March and April 2024. The data analysis was done with SPSS version-26.

## Results

### The socio-demographic characteristics of the study sample

The data represented the general characteristics of 350 participants. In terms of gender, the majority of the participants are female, accounting for 65.4% (229 participants), while males represent 34.6% (121 participants). When it comes to age, 57.4% (201 participants) are 30 years old or younger, and the remaining 42.6% (149 participants) are above 30 years old. In terms of work experience in the field of nursing, 32.0% (112 participants) have more than 5 years of experience, 24.3% (85 participants) have between 3.1 and 5 years of experience, 26.3% (92 participants) have between 1 and 3 years of experience, and 17.4% (61 participants) have less than 1 year of experience. Regarding their professional qualifications, the vast majority hold a Bachelor’s degree, accounting for 92.6% (324 participants). A smaller percentage have a Master’s degree (6.3% or 22 participants), and a very small fraction are PhD scholars or hold a Doctoral Degree (1.1% or 4 participants). Finally, when asked about the type of institute they studied at, 58.9% (206 participants) studied at a private institute, while 41.1% (144 participants) studied at a government institute.

### Knowledge and practise of nursing theories among study participants

The data in Table 2 provides insights into the knowledge and practice of nursing theories. Both knowledge and practice scores were categorized into 3 groups; inadequate (0-3 score), moderately adequate (4-6) and adequate (7-10). In terms of knowledge, 62.0% of the respondents demonstrated adequate knowledge, while 34.0% had moderately adequate knowledge. Only a small fraction, 4.0%, was found to have inadequate knowledge. However, when it comes to the practice of nursing theories, the data tells a different story. A significant 34.6% of the respondents were found to have inadequate practices. Those with moderately adequate practices made up the largest group at 43.7%. Surprisingly, despite the high level of knowledge, only 21.7% of respondents were found to have adequate practices.

**Table 1.** General characteristics of participants (n=350)

| **Variables** | **n (%)** |
| --- | --- |
| **Gender** |  |
| Male | 121 (34.6) |
| Female | 229 (65.4) |
| **Age** |  |
| Less than equal to 30 | 201 (57.4) |
| Above 30 | 149 (42.6) |
| **Designation** |  |
| Nursing Officer | 266 (76.0) |
| Senior Nursing Officer | 66 (18.9) |
| Assistant Nursing Superintendent | 7 (2.0) |
| Deputy Nursing Superintendent | 4 (1.1) |
| Nursing Tutor | 7 (2.0) |
| **Work experience in the field of nursing** |  |
| < 1 year | 61 (17.4) |
| 1-3 years | 92 (26.3) |
| 3.1-5 years | 85 (24.3) |
| > 5 years | 112 (32.0) |
| **Professional Qualification** |  |
| Bachelor degree | 324 (92.6) |
| Master's degree | 22 (6.3) |
| PhD scholar/ Doctoral Degree | 4 (1.1) |
| **Nature of Job** |  |
| Nursing Education | 7 (2.0) |
| Nursing Service | 343 (98.0) |
| **Type of Institute you studied** |  |
| Government | 144 (41.1) |
| Private | 206 (58.9) |

**Table 2.** Response to statements regarding knowledge and utilization of nursing theories [n (%)].

| **SCORE** | **KNOWLEDGE n (%)** | **UTILIZATION n (%)** |
| --- | --- | --- |
| Inadequate | 14 (4.0) | 121 (34.6) |
| Moderately Adequate | 119 (34.0) | 153 (43.7) |
| Adequate | 217 (62.0) | 76 (21.7) |

This suggests a gap between knowledge and practice among the respondents. While most have adequate or moderately adequate knowledge about nursing theories, this knowledge does not seem to translate effectively into practice. This could be due to various factors such as environmental constraints, workload, or lack of reinforcement of training, which could be areas for further investigation and intervention.

### Attitude toward nursing theories among study participants

In the table 3 a significant majority of respondents (93.7%) either strongly agree or agree that there are benefits in patient care if theory-based practice is done. Similarly, 92% of respondents believe that theories have a strong influence and applicability in nursing practice. However, only 57.7% of respondents feel they have a clear understanding of how to use nursing theories in their practice, with 30% uncertain about this.

**Table 3.** Response to statements regarding attitude toward nursing theories [n (%)].

| **ATTITUDE STATEMENT** | **STRONGLY AGREE** | **AGREE** | **UNCERTAIN** | **DISAGREE** | **STRONGLY DISAGREE** |
| --- | --- | --- | --- | --- | --- |
| I believe there are benefits in patient care if theory-based practice is done. | 159 (45.4) | 169 (48.3) | 8 (2.3) | 8 (2.3) | 6 (1.7) |
| Theories have a strong influence and applicability in nursing practice. | 112 (32.0) | 210 (60.0) | 12 (3.4) | 16 (4.6) | 0 (0.0) |
| I have a clear understanding of how to use nursing theories in my practice. | 46 (13.1) | 156 (44.6) | 105 (30.0) | 31 (8.9) | 12 (3.4) |
| Nursing theories differentiate the focus of nursing from that of other professions. | 116 (33.1) | 185 (52.9) | 32 (9.1) | 17 (4.9) | 0 (0.0) |
| I am interested in theory-based practice. | 63 (18.0) | 172 (49.1) | 59 (16.9) | 50 (14.3) | 6 (1.7) |
| Nursing theories help nurses to understand their purpose and role in the health care setting. | 162 (46.3) | 175 (50.0) | 10 (2.9) | 3 (0.9) | 0 (0.0) |
| Theories serve as a rational or scientific reason for nursing interventions. | 152 (43.4) | 161 (46.0) | 21 (6.0) | 16 (4.6) | 0 (0.0) |
| The future of nursing relies on the development of nursing theories. | 106 (30.3) | 182 (52.0) | 44 (12.6) | 0 (0.0) | 18 (5.1) |
| Theories build up positive attitudes among nurses. | 122 (34.9) | 190 (54.3) | 28 (8.0) | 10 (2.9) | 0 (0.0) |
| I am convinced that nursing theory will work and it can be practised | 50 (14.3) | 146 (41.7) | 73 (20.9) | 60 (17.1) | 21 (6.0) |
| I think the application of nursing theories is time-consuming. | 22 (6.3) | 101 (28.9) | 78 (22.3) | 100 (28.6) | 49 (14.0) |
| Nursing theories can be applied in any patient's care despite diagnosis. | 55 (15.7) | 176 (50.3) | 62 (17.7) | 51 (14.6) | 6 (1.7) |
| As a nurse, I feel enthusiastic about using nursing theories. | 40 (11.4) | 121 (34.6) | 82 (23.4) | 91 (26.0) | 16 (4.6) |
| I am ready to incorporate a nursing theory into my practice | 80 (22.9) | 169 (48.3) | 45 (12.9) | 24 (6.9) | 32 (9.1) |

Over 85% of respondents agree that nursing theories differentiate the focus of nursing from that of other professions, and 67.1% are interested in the theory-based practice. Almost all respondents (96.3%) believe that nursing theories help nurses understand their purpose and role in the healthcare setting, and 89.4% agree that theories serve as a rational or scientific reason for nursing interventions.

Looking towards the future, 82.3% of respondents agree that the future of nursing relies on the development of nursing theories, and 89.2% believe that theories build up positive attitudes among nurses. However, only 56% of respondents are convinced that nursing theory will work and can be practised, with 37.2% either disagreeing or strongly disagreeing with this statement.

Interestingly, 35.2% of respondents think the application of nursing theories is time-consuming, with 42.6% either disagreeing or strongly disagreeing. Despite this, 66% of respondents agree that nursing theories can be applied in any patient’s care despite diagnosis. In terms of personal feelings towards using nursing theories, only 46% of respondents feel enthusiastic about this, with 30.6% either disagreeing or strongly disagreeing. However, 71.2% of respondents are ready to incorporate a nursing theory into their practice, indicating a willingness to engage with theory-based practice despite some reservations.

### Levels of knowledge regarding nursing theories among study sample

The data in Table 4 provides an overview of the knowledge levels regarding nursing theories among various participants. The Chi-Square Test values and P-values across all categories indicate that there is no significant difference in the levels of knowledge. This suggests that factors such as designation, age, gender, work experience, professional qualification, nature of the job, and type of institute studied do not significantly influence the levels of knowledge regarding nursing theories among the participants.

**Table 4.**
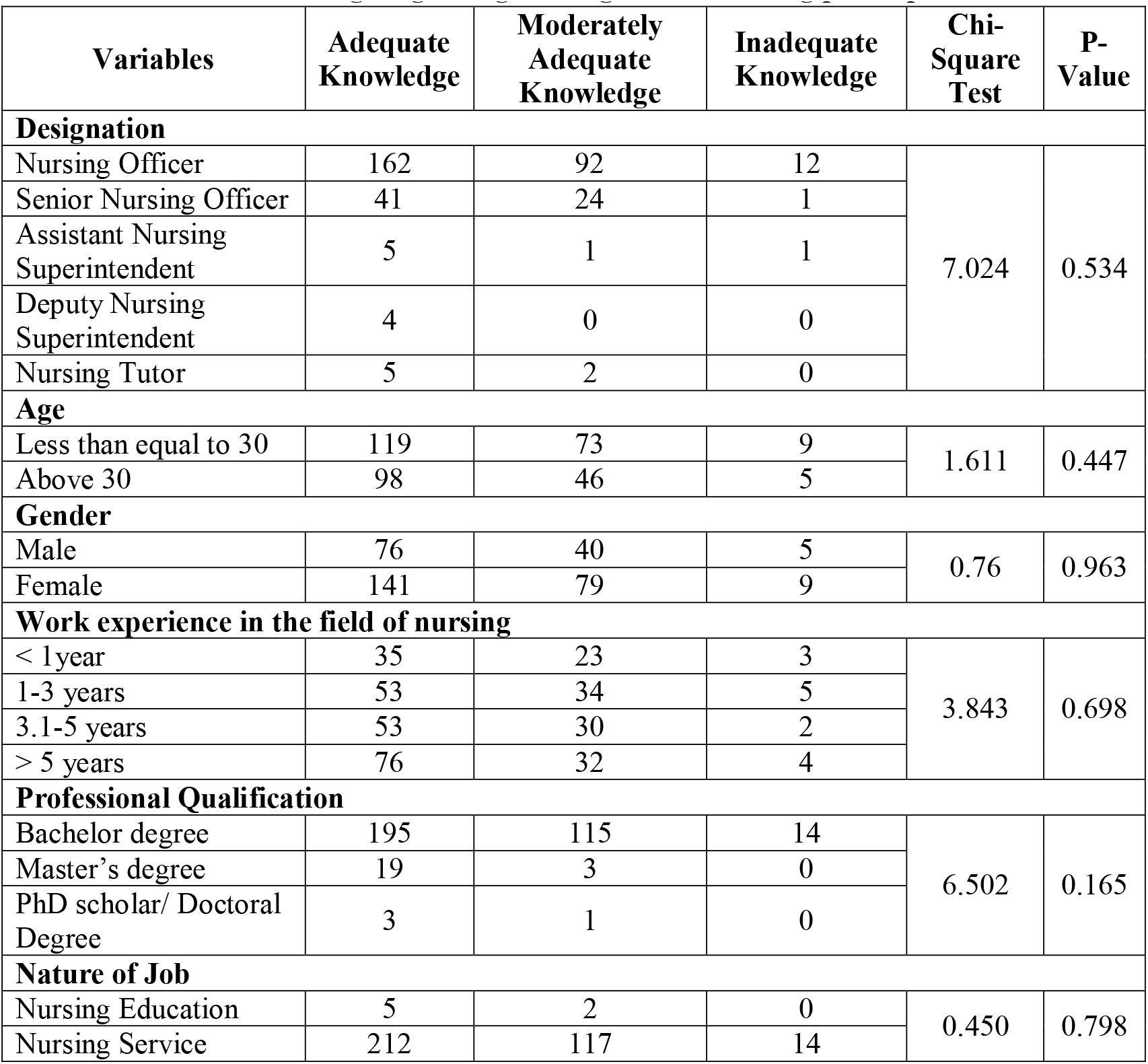
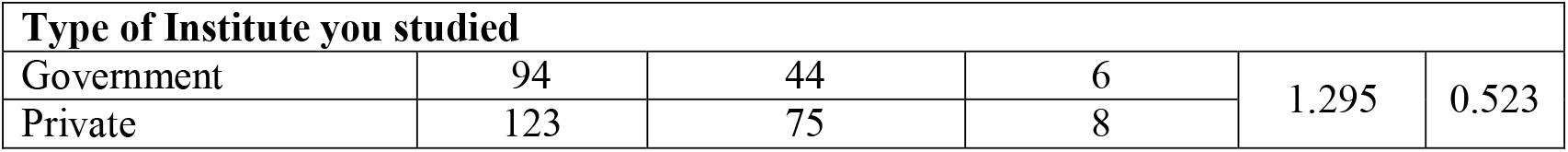
Levels of knowledge regarding nursing theories among participants.

The majority of the participants are Nursing Officers, with 162 of them having adequate knowledge, 92 having moderately adequate knowledge, and 12 having inadequate knowledge. The Chi-square Test value for this group is 7.024 with a P-value of 0.534, suggesting that the difference in knowledge levels across different designations is not statistically significant. In terms of age, participants who are 30 years old or younger seem to have higher knowledge levels, with 119 having adequate knowledge and 73 having moderately adequate knowledge. This is compared to those who are above 30 years of age. When looking at gender, female participants appear to have slightly higher knowledge levels compared to male participants, with 141 females having adequate knowledge and 79 having moderately adequate knowledge. Participants with more than 5 years of experience in the field of nursing have higher levels of adequate knowledge (76) compared to those with less experience. In terms of professional qualifications, participants with a Bachelor’s degree have the highest levels of knowledge, with 195 having adequate knowledge and 115 having moderately adequate knowledge. Participants working in the Nursing Service have higher knowledge levels compared to those in Nursing Education. Additionally, participants who studied in private institutes have slightly higher knowledge levels compared to those who studied in government institutes.

### Levels of practice regarding nursing theories among sample

Table 5 presents a statistical analysis of the levels of practice regarding nursing theories among participants. The factors include designation, age, gender, work experience in the field of nursing, professional qualification, nature of the job, and type of institute studied. The levels of practice are categorized as adequate, moderately adequate, and inadequate. None of the factors show a statistically significant difference in the levels of practice. For instance, the p-value for the factor ‘Designation’ is 0.686, suggesting that the differences in the levels of practice among different designations (Nursing Officer, Senior Nursing Officer, Assistant Nursing Superintendent, Deputy Nursing Superintendent, Nursing Tutor) are not statistically significant. Similarly, the p-value for ‘Age’ is 0.053, ‘Gender’ is 0.660, ‘Work experience in the field of nursing’ is 0.131, ‘Professional Qualification’ is 0.066, ‘Nature of Job’ is 0.447, and ‘Type of Institute you studied’ is 0.961. All these p-values are greater than 0.05, indicating no significant difference in the levels of practice for these factors.

**Table 5.**
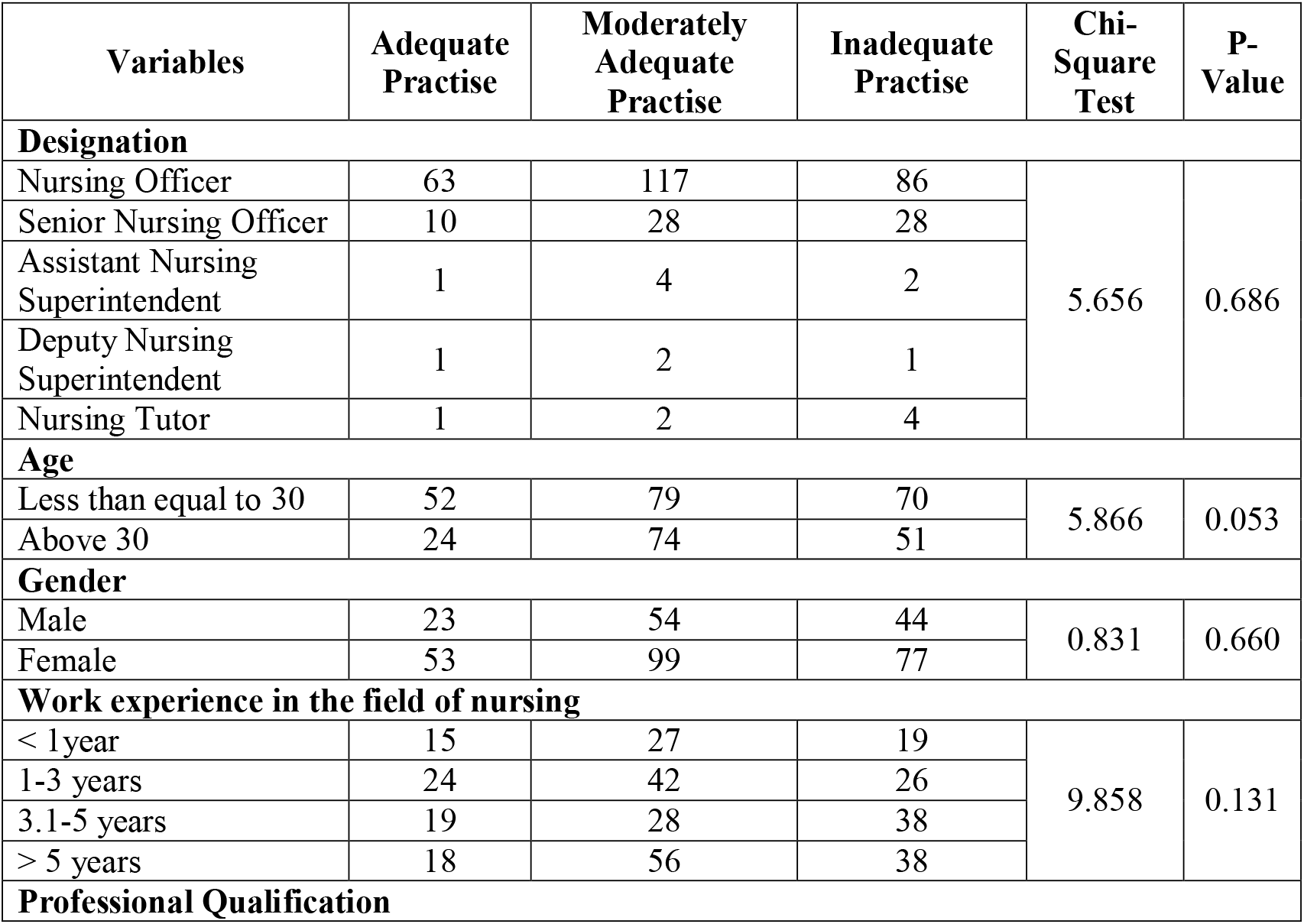
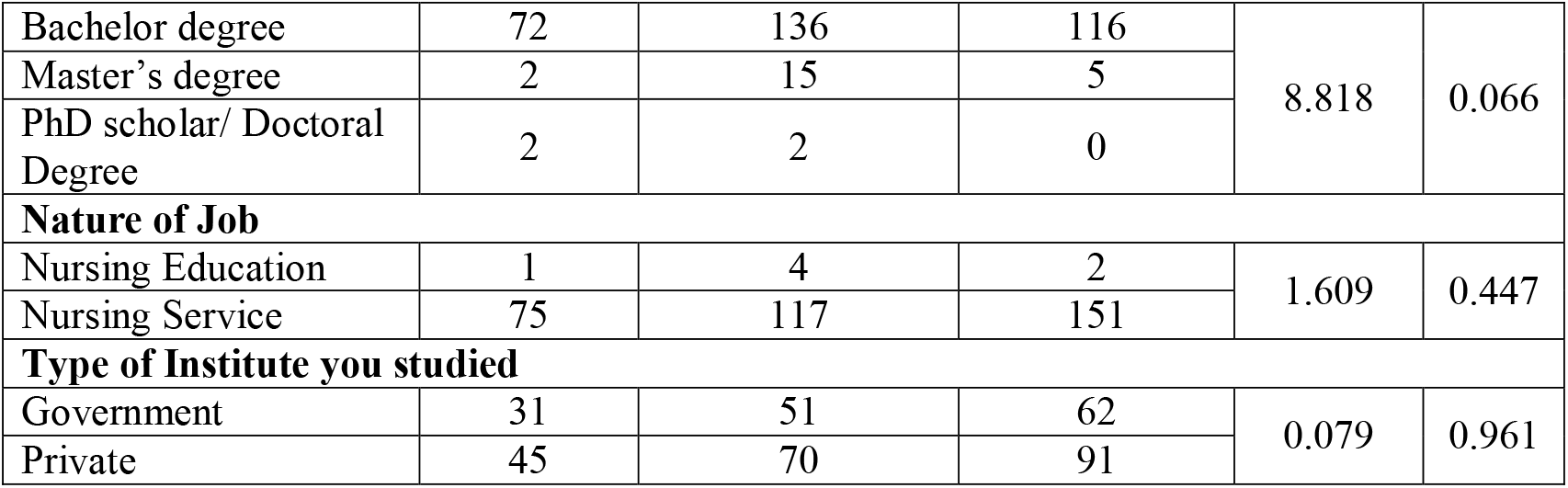
Levels of practice regarding nursing theories among participants.

### Levels of attitude regarding nursing among

The provided Table 6 presents a statistical analysis of level of attitude influencing nursing theory among nursing professionals. The analysis does not show any significant (p-value > 0.05) difference in the mean scores when comparing different designations, age groups, genders, levels of work experience, professional qualifications, and types of institutes, indicating that the observed differences could be due to chance. However, when examining the nature of the job, a significant difference is observed. Specifically, those in Nursing Education have a higher mean score (59.9) compared to those in Nursing Service (53.5). This difference is statistically significant, as indicated by the F-value of 6.783 and the p-value of 0.010. This suggests that the nature of the job, which is Nursing Education, has a positive attitude towards nursing theories.

**Table 6.** Levels of attitude regarding nursing theories among participants.

| Variables | Mean ± SD | SE | F Value | P-Value |
| --- | --- | --- | --- | --- |
| **Designation** |  |  |  |  |
| Nursing Officer | 53.5 ± 6.6 | 0.405 | 3.647 | 0.057 |
| Senior Nursing Officer | 53.1 ± 5.6 | 0.685 | | |
| Assistant Nursing Superintendent | 55.1 ± 3.9 | 1.471 | | |
| Deputy Nursing Superintendent | 54.7 ± 5.1 | 2.562 | | |
| Nursing Tutor | 59.6 ± 8.2 | 3.097 | | |
| **Age** |  |  |  |  |
| Less than equal to 30 | 53.4 ± 6.9 | 0.485 | 0.614 | 0.434 |
| Above 30 | 53.9 ± 5.8 | 0.475 | | |
| **Gender** |  |  |  |  |
| Male | 53.3 ± 5.6 | 0.509 | 0.575 | 0.449 |
| Female | 53.8 ± 6.8 | 0.451 | | |
| **Work experience in the field of nursing** |  |  |  |  |
| < 1year | 54.8 ± 6.6 | 0.839 | 1.745 | 0.187 |
| 1-3 years | 53.8 ± 6.6 | 0.695 | | |
| 3.1-5 years | 52.9 ± 6.6 | 0.716 | | |
| > 5 years | 53.5 ± 6.0 | 0.568 | | |
| **Professional Qualification** |  |  |  |  |
| Bachelor degree | 53.6 ± 6.3 | 0.349 | 1.435 | 0.232 |
| Master's degree | 53.7 ± 8.2 | 1.747 |  |  |
| PhD scholar/ Doctoral Degree | 59.3 ± 6.4 | 2.780 |  |  |
| Nature of Job |  |  |  |  |
| Nursing Education | 59.9 ± 8.2 | 3.097 | 6.783 | 0.010 |
| Nursing Service | 53.5 ± 6.3 | 0.342 |  |  |
| Type of Institute you studied |  |  |  |  |
| Government | 53.9 ± 6.4 | 0.534 | 0.652 | 0.420 |
| Private | 53.4 ± 6.4 | 0.449 |  |  |

## Discussion

To the best of our knowledge, this is the first study to describe nursing personnel’s knowledge, attitude and utilization of nursing theories in India. In order to advance the nursing profession and increase the quality of care, nursing theory application in the clinical setting is highly required. It is necessary to determine, the knowledge, attitude and practice regarding the present state to further understand the barriers and policy changes to be implemented. Since the sub-dimensions of Knowledge, Attitude, and Utilization of nursing personnel towards Nursing Theories Scale were designed to be measured independently, the mean sub-dimension scores were calculated separately. When the results were examined, it was determined that the highest mean score was obtained in the “Attitude” sub-dimension (Table 7).

**Table 7:** Mean and Standard Deviation scores for knowledge, attitude, and practice of nursing theories.

|  | **Knowledge** | **Attitude** | **Practice** |
| --- | --- | --- | --- |
| **Mean** | 2.58 | 53.64 | 1.87 |
| **SD** | 0.570 | 6.433 | 0.740 |

In the present study, the knowledge regarding nursing theories was found to be adequate (62%). But this result was contrary to a cross-sectional study done in two healthcare facilities of Bujumbura, which revealed significantly poor knowledge on nursing theories (75.9%).^5^ The utilization of nursing theories both in the present study was moderately adequate (43.7%) and in the Bujumbara study was poor (83.1%). The reason could be lack of understanding in using nursing theories in practice. Therefore a continuous in-service education can be the only way in overcoming this challenge.

An integrative review on usefulness of nursing theory-guided practice, also revealed an improvement in all of studied outcomes in 26 studies and at least one outcome in nine studies. None of the studies reported that nursing theory-guided interventions as not useful.^6^

The current study also showed a favourable attitude on theory-based practices (49.1%), future of nursing relying on development of nursing theories (52%), theories building up positive attitudes among nurses (54.3%), applicability of theories in any patient care despite medical diagnosis (50.3%) and readiness to incorporate a nursing theory into his/her practice (48.3%). This was similar to a study on attitudes of practicing nurses towards theories in a Canadian setting.^7^ The positive attitude means an improvement in the knowledge levels of nursing personnel could translate into their attitude and improved practice of nursing theories.

### Strengths and Limitations

Despite the good reliability of the tool in the study, caution must be exercised in using the tool as the reliability was checked in a single institute. To our knowledge, there are rarely studies that explore the state of knowledge, attitude and utilization regarding nursing theories. The study was conducted at one tertiary centre, thus limiting generalization. The participants had a lack of interest and acceptability of the topic. They verbalized the fear of being judged.

However, this study has some advantages. It seems to be the first study in India to explore the knowledge, attitudes, and practices of nursing theories among nursing personnel. Findings in this work will help to yield insight into barriers to utilization, provide baseline data for benchmarking, and facilitate planning for nursing theories promotion and improvement by policymakers.

### Implications of the study

Findings in this study identified the level of knowledge, attitudes as well as practices of Nursing Theories in an Indian hospital. These findings provide various stake-holders such as hospital managers and policy makers insight into Nursing Theory practice within Indian hospitals. Stake-holders can use this information to provide the needed resources, training and guidelines for nursing theory based practice among nurses in Indian hospitals to provide good quality patient care.

### Recommendations

Further research can be conducted to assess the barriers on utilization of nursing theories among nursing personnel and can guide policy changes. In-service programs also can be organized to stress importance of theory-based nursing practices.

## Conclusion

Despite nursing theories being taught in the curriculum, less importance is given, which is the reason still nurses lack knowledge on theories. ^8^ Furthermore, examining the state of nursing theory application in healthcare settings may yield insight into barriers to the utilization, provide baseline data for benchmarking and facilitate planning for nursing theories promotion and improvement by policy makers.

## Data availability

The datasets used and/or analysed during the current study available from the corresponding author on reasonable request.

## Acknowledgements

We would like to thank Mrs. Chetna Sahu, Dr. Binu Mathew and Miss. Sanjana for being instrumental in completing the study.

## Declarations

### Funding

This research did not receive any specific grants from any funding agencies.

### Conflicts of Interest

The author(s) declared no specific conflict of interests with respect to the research, authorship and publication of article.

## Notes

### Competing Interest Statement

The authors have declared no competing interest.

### Author Declarations

Prior permission was obtained from Research Review Committee (RRC). CON/AIIMSRPR/RRCN/2023/05 Prior permission was obtained from the Institutional Ethical Committee (IEC). 4273/IEC-AIIMSRPR/2024

